# Estimating the generation interval for COVID-19 based on symptom onset data

**DOI:** 10.1101/2020.03.05.20031815

**Authors:** Ganyani Tapiwa, Kremer Cécile, Chen Dongxuan, Torneri Andrea, Faes Christel, Wallinga Jacco, Hens Niel

**Author notes:** joint first author (Ganyani Tapiwa, Kremer Cécile).

## Abstract

**Background:** Estimating key infectious disease parameters from the COVID-19 outbreak is quintessential for modelling studies and guiding intervention strategies. Whereas different estimates for the incubation period distribution and the serial interval distribution have been reported, estimates of the generation interval for COVID-19 have not been provided.

**Methods:** We used outbreak data from clusters in Singapore and Tianjin, China to estimate the generation interval from symptom onset data while acknowledging uncertainty about the incubation period distribution and the underlying transmission network. From those estimates we obtained the proportions pre-symptomatic transmission and reproduction numbers.

**Results:** The mean generation interval was 5.20 (95%CI 3.78-6.78) days for Singapore and 3.95 (95%CI 3.01-4.91) days for Tianjin, China when relying on a previously reported incubation period with mean 5.2 and SD 2.8 days. The proportion of pre-symptomatic transmission was 48% (95%CI 32-67%) for Singapore and 62% (95%CI 50-76%) for Tianjin, China. Estimates of the reproduction number based on the generation interval distribution were slightly higher than those based on the serial interval distribution.

**Conclusions:** Estimating generation and serial interval distributions from outbreak data requires careful investigation of the underlying transmission network. Detailed contact tracing information is essential for correctly estimating these quantities.

## 1 Introduction

In order to plan intervention strategies aimed at bringing disease outbreaks such as the COVID-19 outbreak under control as well as to monitor disease outbreaks, public health officials depend on insights about key disease transmission parameters which are typically obtained from mathematical or statistical modelling. Examples of key parameters include the reproduction number (average number of infections caused by an infectious individual) and distributions of the generation interval (time between infection events in an infector-infectee pair), serial interval (time between symptom onsets in an infector-infectee pair), and incubation period (time between moment of infection and symptom onset) [1]. Estimates of the reproduction number together with the generation interval distribution can provide insight into the speed with which a disease will spread. On the other hand, estimates of the incubation period distribution can help guide determining appropriate quarantine periods.

As soon as line lists were made available, statistical and mathematical modelling was used to quantify these key epidemiological parameters. Li *et al*. [2] estimated the basic reproduction number (using a renewal equation) to be 2.2 (95% CI 1.4-3.9), the serial interval distribution to have a mean of 7.5 days (95% CI 5.5-19) based on 6 observations, and the incubation period distribution to have a mean of 5.2 days (95% CI 4.1-7.0) based on 10 observations. Other studies estimated the incubation period distribution to have a mean of 6.4 days (95% CI 5.6-7.7) [3], median of 5 days (95% CI 4.0-5.8) [4], mean of 5.2 days (range 1.8-12.4 days) [5], and a mean of 4.8 days (range 2-11 days) [6].

When the incubation period does not change over the course of the epidemic, the expected values of the serial and generation interval distributions are expected to be equal but their variances to be different [7]. It has recently been shown that ignoring the difference between the serial and generation interval can lead to biased estimates of the reproduction number. More specifically, when the serial interval distribution has larger variance than the generation interval distribution, using the serial interval as a proxy for the generation interval will lead to an underestimation of the basic reproduction number *R*. When *R* is underestimated, this may lead to prevention policies that are insufficient to stop disease spread [7].

The most well-known method to estimate the serial interval distribution from line list data is the likelihood-based estimation method proposed by Wallinga and Teunis [8]. In 2012, Hens *et al*. [9] proposed using the Expectation-Maximisation (EM) algorithm to estimate the generation interval distribution from incomplete line list data based on the method by [8] and allowing for auxiliary information to be used in assigning potential infector-infectee pairs. Te Beest et al [10] used a Markov chain Monte Carlo (MCMC) approach as an alternative to the EM-algorithm, to facilitate taking into account uncertainty related to the dates of symptom onset. In this paper, we use an MCMC approach to estimate, next to the serial interval distribution, the generation interval distribution upon specification of the incubation period distribution. We compare the impact of differences amongst previous estimates of the incubation period distribution for COVID-19 and analyse data on clusters of confirmed cases from Singapore (January 21 to February 26) and Tianjin, China (January 14 to February 27).

## 2 Methods

### 2.1 Data

The data used in this paper consist of symptom onset dates and cluster information for confirmed cases in Singapore and Tianjin, China.

As of February 26th, 91 confirmed COVID-19 cases had been reported in Singapore. Detailed information on age, sex, known travel history, time of symptom onset, and known contacts is available for 54 of these cases (link: https://www.moh.gov.sg/news-highlights/, last accessed February 26th). For cases with no infector information available, it is assumed that they could have been infected by any other case within the same cluster. Cases known to be Chinese/Wuhan nationals or known to have been in close contact with a Chinese/Wuhan national are labeled as index cases. All other cases are assumed to have been infected locally.

As of February 27th, 135 confirmed cases had been reported by the Tianjin municipal health commission. Data on these cases are available in official daily reports (link: http://wsjk.tj.gov.cn/col/col87/index.html, last accessed February 27th) and include age, sex, relationship to other known cases, and travel history to risk areas in and outside Hubei province, China. In these data, 114 cases can be traced to one of 16 clusters. The largest cluster consisting of 45 cases can be traced to a shopping mall in Baodi district. Through contact investigations, potential transmission links were identified for cases who had close contacts. Travel history information was used to identify some individuals as import cases. For cases with no infector information available, it is assumed that they could have been infected by any other case within the same cluster.

### 2.2 Model

*For i* = 2, …, *n*, denote *t*_*i*_ the time of infection for individual *i, t*_*v*(*i*)_ the time of infection for the infector of individual *i, δ*_*i*_ the incubation period for individual *i*, and *δ*_*v*(*i*)_ the incubation period for the infector of individual *i*. The serial interval (*Z*_*i*_) for case *i* is a linear combination of latent variables, i.e., *Z*_*i*_ = (*t*_*i*_ + *δ*_*i*_) *−* (*t*_*v*(*i*)_ + *δ*_*v*(*i*)_). Assuming the incubation period is independent of the infection time, *Z*_*i*_ can be rewritten as a convolution of the generation interval for individual *i* and the difference between the incubation period of individual *i* and the incubation period of its infector *v*(*i*) [7], i.e.,

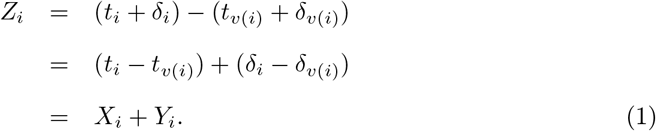

The random variables *X*_*i*_ and *δ*_*i*_ are positive and are both assumed to be independent and identically distributed, i.e., *X*_*i*_ *∼ f* (*x*; Θ_1_) and *δ*_*i*_ *∼ k*(*δ*; Θ_2_), so that *Y*_*i*_ *∼ g*(*y*_*i*_; Θ_2_). Equation (1) implies that both the generation interval and serial interval distributions have the same mean and that the latter has a larger variance and can be negative.

The observed serial interval, *z*_*i*_, can be expressed in terms of the latent variables as *z*_*i*_ = *x*_*i*_ + *y*_*i*_, which implies that, *z*_*i*_ *∼ h*(*z*_*i*_; Θ_1_, Θ_2_). The density function *h*(.) is given by [11],

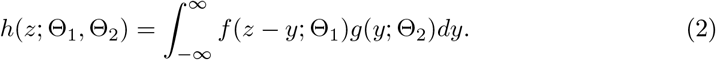

In general, *h*(*z*; Θ_1_, Θ_2_) and *g*(*y*; Θ_2_) have no closed form for arbitrary choices of *f* (*x*; Θ_1_) and *k*(*δ*; Θ_2_). Monte Carlo methods [12] can be used to estimate *h*(*z*; Θ_1_, Θ_2_) as follows,

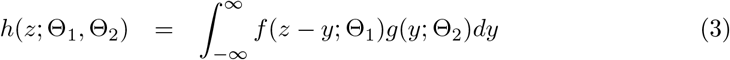

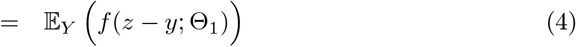

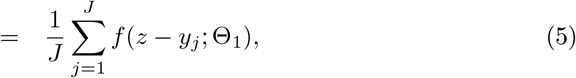

where *y*_*j*_ is the *j*^*th*^ Monte Carlo sample drawn from *g*(*y*; Θ_2_). When all infector-infectee pairs are observed, the likelihood function is given by,

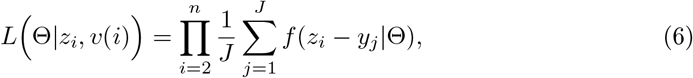

where Θ = *{*Θ_1_, Θ_2_*}* [8]. To account for uncertainty in the transmission links we resort to a Bayesian framework in which missing links are imputed [10] (see Subsection 2.3). The likelihood function is then given by L(Θ, *v*(*i*)^missing^|*z*_*i*_, *v*(*i*))

### 2.3 Estimation

We use the Bayesian method described in te Beest *et al*. [10] for parameter estimation. This method proceeds in two steps. The first step updates the missing links *v*(*i*)^missing^ and the second step updates the parameter vector Θ_1_, i.e., the parameters of the generation interval distribution. We assume that both the generation interval and the incubation period are gamma distributed, i.e., *f* (*x*; Θ_1_) *≡* Γ(*α*_1_, *β*_1_) and *k*(*δ*; Θ_2_) *≡* Γ(*α*_2_, *β*_2_). The parameter vector Θ_2_ is fixed to (*α*_2_ = 3.45, *β*_2_ = 0.66), corresponding to an incubation period with a mean of 5.2 and standard deviation (SD) of 2.8 days [5]. Minimally informative uniform priors are assigned to the parameters of the generation interval distribution, i.e., *α*_1_ *∼ U* (0, 30) and *β*_1_ *∼ U* (0, 20). For cases with multiple potential infectors, the possible links *v*(*i*)^missing^ are assigned equal prior probabilities. The missing links are updated using an independence sampler, whereas Θ_1_ is updated using a random-walk Metropolis-Hastings algorithm with a uniform proposal distribution[12]. We evaluate the posterior distribution using 3 000 000 iterations of which the first 500 000 are discarded as burn-in. Thinning is applied by taking every 200*th* iteration. The serial interval distribution is obtained by simulating 1 000 000 draws from 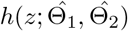. All analyses were performed using R, datasets and code are available on GitHub (https://github.com/cecilekremer/COVID19).

#### 2.3.1 Implications

The proportion of pre-symptomatic transmission is calculated as *p* = *P* (*X*_*i*_ *< δ*_*v*(*i*)_), i.e., pre-symptomatic transmission occurs when the generation interval is shorter than the incubation period of the infector. This proportion was obtained by simulating values from the estimated generation interval and incubation period distributions (assuming a mean incubation time of 5.2 days [5]).

The reproduction number is calculated as 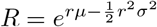, where *r* denotes the exponential growth rate estimated from the initial phase of the outbreak, and *µ* and *σ*^2^ are the mean and variance of the generation or serial interval distribution [13].

Confidence intervals for *p* and *R* are calculated by evaluating *p* and *R* at each iteration of the converged MCMC chain, i.e., at each mean/variance pair of the posterior generation/serial interval distribution. The 95% confidence intervals are given by the 2.5% and 97.5% quantiles of the resulting distributions.

### 2.3.2 Sensitivity analyses

As sensitivity analyses, we investigate the robustness of our estimates of the generation interval distribution to the choice of different incubation period distributions. In particular, we fix Θ_2_ to (*α*_2_ = 7.74, *β*_2_ = 1.21) and (*α*_2_ = 4.36, *β*_2_ = 0.91), corresponding to an incubation period with mean and SD (6.4, 2.3) days [3] and (4.8, 2.6) days [6], respectively.

In our main (i.e., baseline) analyses, missing serial intervals were only allowed to be positive, i.e., the symptom onset time of the infector has to occur before that of the infectee. However, given that pre-symptomatic transmission is possible, this can be deemed an unrealistic assumption. Therefore, we assess the impact of allowing for negative serial intervals on our estimates of the generation interval distribution.

To further assess the robustness of the estimated generation interval distribution, for each dataset, we fit the model to data from the largest cluster. In the Tianjin dataset, the largest cluster is the shopping mall cluster consisting of 45 cases. In the Singapore dataset this is the Grace Assembly of God cluster consisting of 25 cases.

## 3 Results

### 3.1 Estimates of key epidemiological parameters

Table 1 shows parameter estimates of the generation and serial interval distributions for each dataset, assuming an incubation period with mean 5.2 and SD 2.8 days. The mean generation time is estimated to be 5.2 days (95%CI, 3.78 - 6.78) for the Singapore data, and 3.95 days (95%CI, 3.01 - 4.91) for the Tianjin data. As expected, the estimated means of the generation interval and serial interval distributions are approximately equal but the latter has a larger variance.

**Table 1:** Parameter estimates and credible intervals of generation and serial interval distributions for Singapore and Tianjin datasets. The estimates are obtained using reported information on infector-infectee pairs and assuming an incubation period with mean 5.2 and SD 2.8 days. GI: generation interval; SI: serial interval.

| Data | Scenario | Interval | Estimate (95% credible interval) |  |
| --- | --- | --- | --- | --- |
|  |  |  | Mean | Standard deviation |
| Singapore | Baseline | GI | 5.20 (3.78, 6.78) | 1.72 (0.91, 3.93) |
|  |  | SI | 5.21 (-3.35, 13.94) | 4.32 (4.06, 5.58) |
| Tianjin<br>(China) | Baseline | GI | 3.95 (3.01, 4.91) | 1.51 (0.74, 2.97) |
|  |  | SI | 3.95 (-4.47, 12.51) | 4.24 (4.03, 4.95) |

### 3.2 Sensitivity analyses

Table 2 shows parameter estimates of the generation and serial interval distributions for each dataset, assuming incubation periods with mean and SD (6.4, 2.3) days and (4.8, 2.6) days. The parameter estimates are fairly robust to the specified incubation period distribution, with mean generation times about 5 days for Singapore and 4 days for Tianjin.

**Table 2:** Parameter estimates and credible intervals of generation and serial interval distributions for Singapore and Tianjin datasets. The estimates are obtained under different assumptions for the incubation period, under the baseline scenario where missing SI are only allowed to be positive. GI: generation interval; SI: serial interval

| Data | Incubation period | Interval | Estimate (95% credible interval) |  |
| --- | --- | --- | --- | --- |
|  |  |  | Mean | Standard deviation |
| Singapore | Mean 6.4 SD 2.3 | GI | 5.29 (3.89, 6.77) | 2.08 (0.97, 4.07) |
|  |  | SI | 5.29 (-2.13, 13.16) | 3.86 (3.40, 5.21) |
|  | Mean 4.8 SD 2.6 | GI | 5.19 (3.82, 6.74) | 1.77 (0.91, 4.11) |
|  |  | SI | 5.19 (-2.86, 13.45) | 4.08 (3.79, 5.51) |
| Tianjin<br>(China) | Mean 6.4 SD 2.3 | GI | 4.02 (3.11, 5.00) | 2.29 (1.02, 3.80) |
|  |  | SI | 4.02 (-4.83, 13.45) | 3.98 (3.41, 5.00) |
|  | Mean 4.8 SD 2.6 | GI | 3.95 (3.05, 4.93) | 1.75 (0.77, 3.35) |
|  |  | SI | 3.95 (-4.60, 12.73) | 4.07 (3.76, 4.97) |

Table 3 shows parameter estimates of the generation and serial interval distributions obtained when allowing for negative serial intervals in case there is no known infector. Compared to baseline analyses (Table 1), estimates of the mean generation time are smaller when allowing for negative serial intervals. The mean generation time is 3.86 days for Singapore and 2.90 days for Tianjin.

**Table 3:** Parameter estimates and credible intervals of generation and serial interval distributions for Singapore and Tianjin datasets when allowing serial intervals to be negative and assuming an incubation period with mean 5.2 and SD 2.8 days. GI: generation interval; SI: serial interval

| Data | Scenario | Interval | Estimate (95% credible interval) |  |
| --- | --- | --- | --- | --- |
|  |  |  | Mean | Standard deviation |
| Singapore | Allowing for all possible negative SI | GI | 3.86 (2.22, 5.60) | 2.65 (0.87, 5.43) |
|  |  | SI | 3.86 (-5.15, 13.88) | 4.76 (4.05, 6.72) |
| Tianjin<br>(China) | Allowing for all possible negative SI | GI | 2.90 (1.85, 4.12) | 2.86 (1.37, 5.04) |
|  |  | SI | 2.90 (-6.12, 13.47) | 4.88 (4.19, 6.41) |

Table 4 shows parameter estimates obtained when we fit the model to data from the largest cluster. We only show results for the Tianjin dataset, as for the Singapore data there were too few cases (*n*=25) and the MCMC chain did not converge. When allowing only positive serial intervals for cases with no known infector, the mean generation time is estimated to be 3.50 days. On the other hand, when allowing for negative serial intervals, it is estimated to be 2.57 days.

**Table 4:** Parameter estimates and credible intervals of generation and serial interval distributions for the largest cluster in the Tianjin dataset. The estimates are obtained under different scenarios for the serial interval, assuming an incubation period with mean 5.2 and SD 2.8 days. GI: generation interval; SI: serial interval

| Data | Scenario | Interval | Estimate (95% credible interval) |  |
| --- | --- | --- | --- | --- |
|  |  |  | Mean | Standard deviation |
| Tianjin<br>(China) | Baseline | GI | 3.50 (2.10, 5.03) | 1.70 (0.65, 4.10) |
|  |  | SI | 3.50 (-5.02, 12.25) | 4.31 (4.01, 5.70) |
|  | Allowing for all possible negative SI | GI | 2.57 (1.14, 4.30) | 2.58 (0.68, 6.11) |
|  |  | SI | 2.57 (-6.28, 12.70) | 4.72 (4.02, 7.28) |

### 3.3 Implications

Table 5 shows the proportions of pre-symptomatic transmission and reproduction numbers for each dataset. Pre-symptomatic transmission is higher when allowing for negative serial intervals for cases with no known infector. The reproduction number is lower when estimated using the serial interval compared to when using the generation interval.

**Table 5:** Proportion of pre-symptomatic transmission (*p*) and reproduction number *R* estimated using GI or SI, assuming an incubation period with mean 5.2 and SD 2.8 days. GI: generation interval; SI: serial interval

| Data | Scenario | Interval | Estimate (95% credible interval) |  |
| --- | --- | --- | --- | --- |
| | | | $p$ | $R$ |
| Singapore | Baseline | GI | 0.48 (0.32, 0.67) | 1.27 (1.19, 1.36) |
|  |  | SI | - | 1.25 (1.17, 1.34) |
|  | Allowing for all possible negative SI | GI | 0.66 (0.45, 0.84) | 1.19 (1.10, 1.28) |
|  |  | SI | - | 1.17 (1.08, 1.26) |
| Tianjin<br>(China) | Baseline | GI | 0.62 (0.50, 0.76) | 1.59 (1.42, 1.78) |
|  |  | SI | - | 1.41 (1.26, 1.58) |
|  | Allowing for all possible negative SI | GI | 0.77 (0.65, 0.87) | 1.32 (1.18, 1.51) |
|  |  | SI | - | 1.17 (1.05, 1.34) |

## 4 Discussion

We estimated the generation time to have a mean of 5.20 (95%CI 3.78-6.78) days and a standard deviation of 1.72 (95%CI 0.91-3.93) days for the Singapore data, and a mean of 3.95 (95%CI 3.01-4.91) days with a standard deviation of 1.51 (95%CI 0.74-2.97) days for the Tianjin data. These mean estimates increase only slightly when increasing the mean incubation period. For the Singapore data, allowing the serial interval to be negative decreases the estimated mean generation time from 5.20 days, when restricting missing serial intervals to be positive, to 3.86 (95%CI 2.22-5.60) days when allowing them to be negative. For the Tianjin data, the baseline estimate of the mean generation time (3.95 days) is about the same than when allowing serial intervals to be negative in the Singapore data. However, in the Tianjin data there were already some negative serial intervals among the reported links, which may explain this lower estimate. When allowing for negative serial intervals in the Tianjin data, the mean generation time decreased to 2.90 (1.85-4.12) days. These are the first estimates of the generation interval for COVID-19. Sensitivity analyses show that the assumptions made about the incubation period have only moderate impact on the results. On the other hand, assumptions made about the underlying transmission network (e.g. acknowledging possibly negative serial intervals) have a large impact on our results.

As expected, the proportion of pre-symptomatic transmission increases from 48% (95%CI 32-67%) in the baseline scenario to 66% (95%CI 45-84%) when allowing for negative serial intervals, for the Singapore data, and from 62% (95%CI 50-76%) to 77% (95%CI 65-87%) for the Tianjin data. Hence, a substantial proportion of transmission appears to occur before symptom onset, which is an important point to consider when planning intervention strategies. We also estimated *R*_0_, solely to illustrate the bias that occurs when using the serial interval as a proxy for the generation interval [7]. Whereas the impact was limited for our analyses, estimates based on the generation interval are larger and should be preferred to inform intervention policies. Indeed, as expected, the reproduction number was underestimated when using the serial interval distribution which is more variable than the generation interval distribution.

Tindale *et al*. [14] recently estimated the mean serial interval for COVID-19 to be 4.56 (95%CI 2.69 - 6.42) days for Singapore and 4.22 (95%CI 3.43 - 5.01) days for Tianjin. Although these estimates are different from the ones we report, they fall within the uncertainty ranges we obtained. An important advantage of our method is that we are able to infer the generation interval distribution while allowing serial intervals to be negative. Our estimates of *R* are smaller than the ones reported by Tindale *et al*. [14], because we use a different estimate of the growth rate *r* (0.04 for Singapore and 0.12 for Tianjin as obtained from the initial exponential growth phase in each dataset, compared to 0.15 used by [14]).

Another advantage of our method is that we can derive a proper variance estimate for the generation interval, in contrast to using a too large variance estimate that is obtained when using the serial interval as a proxy for the generation interval. Furthermore, in theory we do not need to condition on the order of symptom onset times. However, when the data does not provide sufficient information on directionality of transmission, this lack of auxiliary information may cause problems for estimation.

This study does have some limitations. First, we rely on previous estimates for the incubation period. However, sensitivity analyses show that changing the incubation period distribution does not have a big impact on our estimates of the generation interval distribution. Second, we do not account for incomplete or possible changes in reporting over the course of the epidemic. Third, we do not acknowledge changes in contact processes and thus behavioral change, which could shape realised generation interval distributions as well as serial interval distributions (unpublished work). Fourth, we do not account for contraction of the generation interval because of depletion of susceptibles. Future work should take into account these shortcomings.

Infection control for the COVID-19 epidemic relies on case-based measures such as finding cases and tracing contacts. A variable that determines how effective these case-based measures are is the proportion of pre-symptomatic transmission. Our estimates of this proportion are high, ranging from 0.48 to 0.77. This implies that the effectiveness of case finding and contact tracing in preventing COVID-19 infections will be considerably smaller compared to the effectiveness in preventing SARS or MERS infections, where pre-symptomatic transmission did not play an important role (see e.g. [15]). It is unlikely that these measures alone will suffice to control the COVID-19 epidemic. Additional measures, such as social distancing, are required.

## Data Availability

All data are available on GitHub.

https://github.com/cecilekremer/COVID19

## Acknowledgements

NH acknowledges funding from the European Research Council (ERC) under the European Unionâs Horizon 2020 research and innovation programme (grant agreement 682540 - TransMID). CF, NH and JW acknowledge funding from the European Union’s SC1-PHE-CORONAVIRUS-2020 programme.

